# *Toxoplasma* Infection and Its Sequential Impact on Physical Health, Stress, and Anxiety: A Large Cross-Sectional Study Testing the Stress-Coping Hypothesis

**DOI:** 10.1101/2024.10.21.24315879

**Authors:** Jaroslav Flegr, Ashkan Latifi, Šárka Kaňková

## Abstract

**Introduction:** Latent toxoplasmosis, affecting approximately one-third of people worldwide, was once thought to be asymptomatic. However, studies in the last three decades have revealed that it can cause significant psychological and behavioral changes in humans. The observation that the behavioral impacts of toxoplasmosis manifest in opposite directions in men and women has led to the development of the Stress-Coping Hypothesis. This hypothesis posits that health degradation from toxoplasmosis results in chronic stress, with gender-specific coping strategies explaining the divergent behavioral responses observed between men and women.

**Methods:** This study, conducted on 1,768 individuals who had previously been tested for toxoplasmosis or borreliosis, sought to examine this hypothesis through a survey that included the Perceived Stress Scale and the State-Trait Anxiety Inventory.

**Results:** Confirmed poorer health, higher stress, and anxiety levels among *Toxoplasma*-infected participants. Path analysis showed that toxoplasmosis directly negatively impacts physical health, which in turn directly increases stress and anxiety among infected individuals, thereby negatively affecting cognitive performance. This pattern was not seen with borreliosis, serving as a negative control, underscoring the unique impact of toxoplasmosis on human physical health, well-being, and cognition.

**Conclusions:** Our findings strongly suggest that the cognitive impairments associated with toxoplasmosis are primarily side effects of chronic stress resulting from the compromised health of infected individuals.

**Key Points:** Chronic stress mediates the negative effects of toxoplasmosis on cognition, stemming from the impaired health of infected individuals.

This effect was not observed with the negative control, borreliosis.

Our results support the side effects hypothesis and contradict the parasite manipulation hypothesis.

## 1. Introduction

Toxoplasmosis is a zoonotic parasitic infection caused by the obligate intracellular protozoan *Toxoplasma gondii.* It is estimated that currently about one-third of the population in both developed and developing countries is infected, with the infection, characterized in its later stage by the presence of dormant stages in the bodies of infected individuals, likely being lifelong (Pappas, Roussos, & Falagas, 2009; Tenter, Heckeroth, & Weiss, 2000). To undergo sexual reproduction, *Toxoplasma gondii* must infect the epithelial cells of the small intestine in its definitive host, a feline. It is within these intestinal epithelial cells that the parasite undergoes sexual reproduction, leading to the production of oocysts. Once this process is complete, the oocysts are expelled into the environment through the host’s feces, making them available to infect intermediate hosts. These are mostly rodents, but almost every type of warm-blooded vertebrates that happen to ingest the oocytes can get infected. In the bodies of intermediate hosts, sporozoites released from ingested oocysts invade cells and transform into tachyzoites, which then begin to rapidly asexually reproduce in various tissues and organs, including the brain, heart, and skeletal muscles. Over time, this process leads to the formation of tissue cysts, the transformed host cells, containing bradyzoites, marking the onset of lifelong latent toxoplasmosis.

*Toxoplasma* is transmitted vertically, from the mother in the acute stage of infection to the fetus, and horizontally, through ingestion of oocytes or through consumption of the infected intermediate host by the definitive host or other intermediate hosts. The former, congenital toxoplasmosis, leads to far more severe complications, including developmental defects and miscarriages of infected fetuses (Pappas et al., 2009; Tenter et al., 2000). Although horizontal transmission of *Toxoplasma* and the resultant postnatally acquired toxoplasmosis were long considered asymptomatic in immunocompetent hosts, studies in the last approximately 30 years using animal and human models have uncovered impacts on the infected intermediate host in several aspects. Among these impacts are behavioral changes, which are most probably the result of manipulations by the parasite, see (Abdulai-Saiku, Tong, & Vyas, 2017; Saadatnia & Golkar, 2012; Webster, Brunton, & Macdonald, 1994; Wohlfert, Blader, & Wilson, 2017).

These manipulations enhance the likelihood of an intermediate host being consumed by a definitive host (Pardo Gil et al., 2023). Notably, one such manipulation involves the intermediate host developing an attraction to the urine odor of the definitive host, effectively drawing it closer to its natural predator. This phenomenon has been observed in *Toxoplasma*- infected rodents (Berdoy, Webster, & Macdonald, 2000), chimpanzees (Poirotte et al., 2016), and humans (Flegr, Lenochová, Hodný, & Vondrová, 2011). In modern humans, who are no longer primary prey for felines, the effects of infection with this parasite have also been observed in various other domains, such as personality (Flegr, Ullmann, & Toman, 2023; Flegr, Zitkova, Kodym, & Frynta, 1996), psychopathology (Flegr & Horáček, 2020; Rahiminezhad, Latifi, Rostami, & Farahani, 2024), cognition (de Haan, Sutterland, Schotborgh, Schirmbeck, & de Haan, 2021), and behavior (Lerner, Alkærsig, Fitza, Lomberg, & Johnson, 2020; Lindová et al., 2006).

Studies done on the relationship between latent toxoplasmosis and personality change showed that the effects of *Toxoplasma* infection on personality traits were often opposite in infected men compared with infected women. Examples of such traits include increased cooperativeness, trustfulness, superego strength, and extroversion in *Toxoplasma*-infected women, contrasted with a decrease in these traits in *Toxoplasma*-infected men (Flegr & Hrdý, 1994; Flegr et al., 1996; Lindová et al., 2006). As a potential explanation for this observation, researchers have proposed a psychological rationale focusing on the distinct coping strategies employed by men and women in response to prolonged stress caused by infection (Lindová et al., 2010; Lindová et al., 2006). It is known that whilst men are more inclined to employ individualistic, rational, and detachment coping strategies, women are more likely to implement emotional and avoidance-based coping strategies, and also resort to social support when experiencing stress (Kelly, Tyrka, Price, & Carpenter, 2008; Matud, 2004). In the study by Lindová et al. (Lindová et al., 2006), the health decline caused by prolonged stress from infection is suggested to lead *Toxoplasma*-infected individuals to adopt gender-specific coping strategies. These strategies might explain the observed divergent personality traits between infected males and females.

Latent toxoplasmosis is indeed found to negatively affect the physical health of the infected individuals (Flegr & Escudero, 2016); also see (Flegr, Prandota, Sovickova, & Israili, 2014).

However, few if any studies have addressed the empirical question of the heightened levels of stress in *Toxoplasma*-infected subjects compared with uninfected controls. A typical behavioral symptom of chronic stress is anxiety, (Coppola & Spector, 2009; Endler & Parker, 1990; Hussenoeder et al., 2022; Qin et al., 2015; Smith, Metzker, Waite, & Gerrity, 2015).

Consequently, we can predict that higher levels of anxiety would be observed in infected subjects compared to noninfected ones. In line with that, laboratory mice in the latent stage of *Toxoplasma* infection exhibit anxiety-like behavior (Bay-Richter, Petersen, Liebenberg, Elfving, & Wegener, 2019), and an increased incidence of Generalized Anxiety Disorder has been reported in *Toxoplasma*-infected humans (Akaltun, Kara, & Kara, 2018; Flegr & Horáček, 2020; Suvisaari, Torniainen-Holm, Lindgren, Harkanen, & Yolken, 2017). Surprisingly, data demonstrating an association between latent toxoplasmosis and increased anxiety as a trait in the nonclinical population are scarce. The sole study indicating increased anxiety (along with depression and obsession) in *Toxoplasma*-infected individuals, conducted on a population of about 3,600 subjects, did not measure personality traits anxiety with psychological questionnaires but relied on participants’ subjective ratings of the intensity of suffering from specific neuropsychiatric symptoms (depression, mania, phobia, anxiety, and obsessions) on a 100-point scale (Flegr & Horáček, 2020).

Another zoonotic disease, although much less comprehensively studied than toxoplasmosis in relation to potential infection-induced behavioral changes, is borreliosis. Borreliosis is caused by the bacterium *Borrelia burgdorferi* (Nadelman & Wormser, 1998). *Borrelia* is transmitted through tick bites and may persist for years in an infected human’s brain, heart, liver, and kidney, even after extensive antibiotic therapy (Sapi et al., 2019), suggesting its potential for a life-long latency. The seroprevalence of borreliosis is reported to be 13.6% in Western Europe, 11.1% in Eastern Europe, 4.2% in Northern Europe, and 3.9% in Southern Europe by a meta-analysis of the related studies which used two-tier testing from 2005 to 2020 (Burn et al., 2023). While latent borreliosis, unlike the much rarer neuroborreliosis, was historically considered to be almost asymptomatic, it is essential to acknowledge recent findings from two studies indicating that borreliosis can lead to deteriorated health in infected individuals (Flegr & Horáček, 2018; Flegr et al., 2023). Given the nature of its life cycle, it is not expected that *Borrelia* would alter the behavior or psychological state of infected individuals. Moreover, it was demonstrated that, unlike latent toxoplasmosis, latent borreliosis manifests only in poorer physical health, not in mental health (Flegr & Horáček, 2018; Flegr et al., 2023).

The general aim of the study was to investigate the basic premise of the validity of the Stress-Coping Hypothesis, specifically examining the level of perceived stress in *Toxoplasma*-infected subjects. The secondary aim was to use path analysis to distinguish the direct effect of toxoplasmosis on stress and anxiety from the effects mediated by impaired physical health.

Data was gathered from 1,768 nonclinical internet users who had been laboratory tested for toxoplasmosis, using an electronic survey that included the Perceived Stress Scale and the State-Trait Anxiety Inventory. To ensure that any identified association between stress and infection wasn’t merely a byproduct of the survey method, that is, the potential tendency of individuals with heightened health awareness to report both poorer health and infection, the reported presence of *Borrelia* infection was utilized as a form of negative control.

## 2. Material and Methods

### 2.1. Participants

Participants in the study were recruited using a Facebook-based snowball sampling method (S. Kaňková, Flegr, & Calda, 2015). Invitations to participate in an online survey, focusing on the well-being and discomfort of Czech residents during the COVID-19 pandemic, were posted on several Facebook pages. At the end of the survey, participants were encouraged to share and recommend the questionnaire to their friends, leading to a broad and diverse range of respondents. The introductory page of the approximately 10-minute survey informed participants about the topics covered, including basic demographic data, personality, life satisfaction, and opinions on the current situation. They were also assured of anonymity, the exclusive research purpose of the collected data, its confidentiality, the voluntary nature of participation, and the option to withdraw at any time by closing the webpage. Informed consent was obtained by participants clicking a button on the page. Participants were not financially compensated for their participation in the study. Instead, they received their personal results from the STAI-X2 and PSS questionnaires, along with histograms showing the distribution of these results among participants from the previous similar study. The survey, part of the project ‘Influence of Biological, Socioeconomic, and Psychological Factors on People’s Behavior and Attitudes in Relation to Epidemiology’ was approved by the Institutional Review Board of the Faculty of Science, Charles University and conducted following all relevant guidelines and regulations. The final version was launched from August 28, 2020, through the end of the year, with most of the 7,214 respondents completing it by the end of October 2020.

### 2.2. Questionnaires

Two standardized questionnaires were employed for collecting data on anxiety and stress in our sample. State Trait Anxiety Inventory X-2 (STAI-X2), re-standardized by (Heretik, Ritomský, Novotný, Heretik, & Pečeňák, 2009) in an adult Slovak population, was implemented to evaluate the anxiety trait in the participants. Heretik et al. reported alpha reliability coefficients of 0.85 through 0.88 for this questionnaire. STAI-X2 is composed of 20 items scaled on a 4-point Likert scale (almost never, sometimes, often, almost always). Higher scores on STAI-X2 indicate higher levels of anxiety trait. On the STAI-X2, the lowest possible score is 20, and the highest possible score is 80. Stress was measured via implementation of the Perceived Stress Scale (PSS), which nests 10 items scaled on a 5-point Likert scale (never, almost never, sometimes, quite often, very often). On the PSS, 0 is the lowest possible score, and 40 is the highest possible score, with the higher scores denoting higher levels of perceived stress. The PSS was originally developed by (Cohen, Kamarck, & Mermelstein, 1983), who reported alpha reliability coefficients of 0.84, 0.85, and 0.86 in the three samples that they used in their studies. We also assessed the reliability of these questionnaires in our sample for all participants, and also for men and women separately. The reliability coefficients of McDonald’s omega total and Cronbach’s alpha for STAI-X2 for all participants were 0.94 and 0.93, respectively. They proved 0.94 and 0.93 for women, and 0.93 and 0.92 for men on this questionnaire, respectively. We also calculated the McDonald’s omega total and Cronbach’s alpha for PSS, which resulted in the coefficients of 0.90 and 0.88 for all participants, 0.90 and 0.88 for women, and 0.89 and 0.87 for men, respectively. Overall, these results demonstrated acceptable reliabilities for these instruments in our study. In addition, STAI-X2 exhibited higher reliability coefficients for all participants, men, and women when compared with PSS. On the whole, STAI-X2 and PSS had slightly higher reliability in the women sample compared to the men sample.

In the final part of the questionnaire, the participants were asked whether or not they had been laboratory tested for toxoplasmosis or borreliosis and were required to self-report their infection status based on the laboratory results. They were reminded that toxoplasmosis is a parasite of cats that is especially dangerous for pregnant women, and also that borreliosis is a tick-borne bacterial disease transmitted through the bite of ticks. Accordingly, in this regard, they could choose one of the three options available to them (1) positive - I was tested and the result of the test was positive, 2) negative - I was tested and the result of the test was negative, or 3) I do not know, I am not sure). Our participants also self-rated their physical health using a scale of 0 to 100, indicating an absolute lack of physical health to an absolute state of health, respectively. Participants also disclosed their sex assigned at birth, their age, and the size of their place of residence, which was rated on a 7-point ordinal scale. The overarching aim of the entire project was to explore the effects of the pandemic and lockdowns on the well-being of the general Czech population; therefore, the questionnaire included numerous questions unrelated to the current study.

### 2.3. Statistical Analysis

We conducted the Shapiro-Wilk test and examined data distribution plots to assess the normality assumption. Upon identifying violations of this assumption, we applied non-parametric tests to address these issues. Nonetheless, comparisons between parametric and non-parametric tests revealed nearly identical results. Categorical data, namely sex and infection status, were cross-tabulated, and Chi-square tests were used to uncover their potential associations.

We conducted both multivariate and univariate analyses of covariance (MANCOVA and ANCOVA) using Type III sum of squares to investigate the effects of infection and sex, as well as their interactions, on anxiety and stress, with age as a control. This approach allowed us to accurately assess each factor’s unique contribution by adjusting for the variance explained by other variables in the model, ensuring robust analysis in potentially unbalanced designs or when multiple covariates are involved. We also considered the size of the place of living as a potential confounder. However, AIC comparisons showing minimal difference between simpler and complex models led us to exclude the living place size from our final analysis. The effects of sex and infection on age, physical health, anxiety, and stress were further analyzed using univariate non-parametric Wilcoxon rank-sum tests and multivariate non-parametric Kendall’s Tau partial correlation tests, with adjustments for age and, where necessary, for sex and physical health.

Path analysis was employed for an in-depth exploration of the relationships among various variables. This method allowed for the differentiation between direct and indirect effects of the primary interest variables, specifically toxoplasmosis and borreliosis. It further facilitated the description and distinction of the mediating and confounding effects of physical health and age.

We used R (version 4.3.0) for doing descriptive statistics, Wilcoxon rank-sum test, ANCOVA, and path analysis (via packages lavaan and semPlot). Non-parametric Kendall Tau partial correlations were conducted using Explorer 1.0 (Flegr & Flegr, 2021). We also used SPSS (version 26) for MANCOVA modeling.

All data are available at figshare https://doi.org/10.6084/m9.figshare.25188788.v1.

**Technical notes:** (1) Except for the section Discussion, we use the terms “toxoplasmosis” and “borreliosis“ as a shorthand for scoring *Toxoplasma*- or *Borrelia*-seropositive in standard IgG serological tests. (2) Throughout the manuscript, we use the technical term “effect” in a sense used in statistics, i.e., as referring to the difference between the true population parameter and the null hypothesis value. The Discussion section is the only place where we discriminate between cause and effect and use “effect” in its nontechnical sense.

## 3. Results

### 3.1. Descriptive Statistics

The initial dataset comprised 7,214 individuals. For this study, we focused on the 1,768 subjects who reported a test result for toxoplasmosis or borreliosis. We divided them into three groups: the Toxoplasmosis Test Group (698 subjects tested for toxoplasmosis: 172 men, 526 women), the Borreliosis Test Group (1,584 subjects tested for borreliosis: 650 men, 934 women), and the Dual Test Group (514 subjects tested for both pathogens: 148 men, 366 women). The Dual Test Group was utilized only for the MANCOVA and ANCOVA analyses that addressed both infections in a single model to assess their potential interaction. Descriptive statistics for the overall dataset are illustrated in Figure 1, while the descriptive statistics for the aforementioned groups are detailed in Table 1.

**Figure 1.**
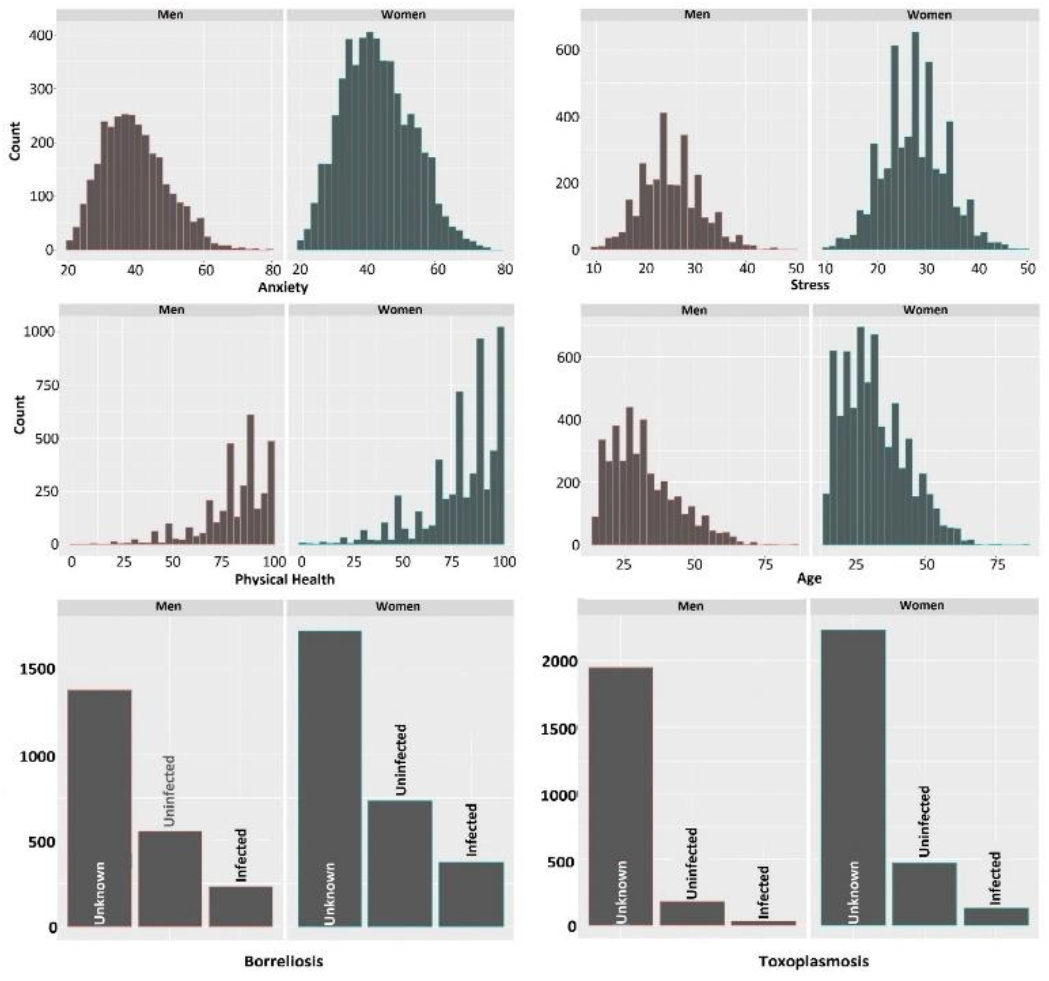
Data distribution of physical health, anxiety, stress, borreliosis, toxoplasmosis, and age for men and women

**Table 1.**
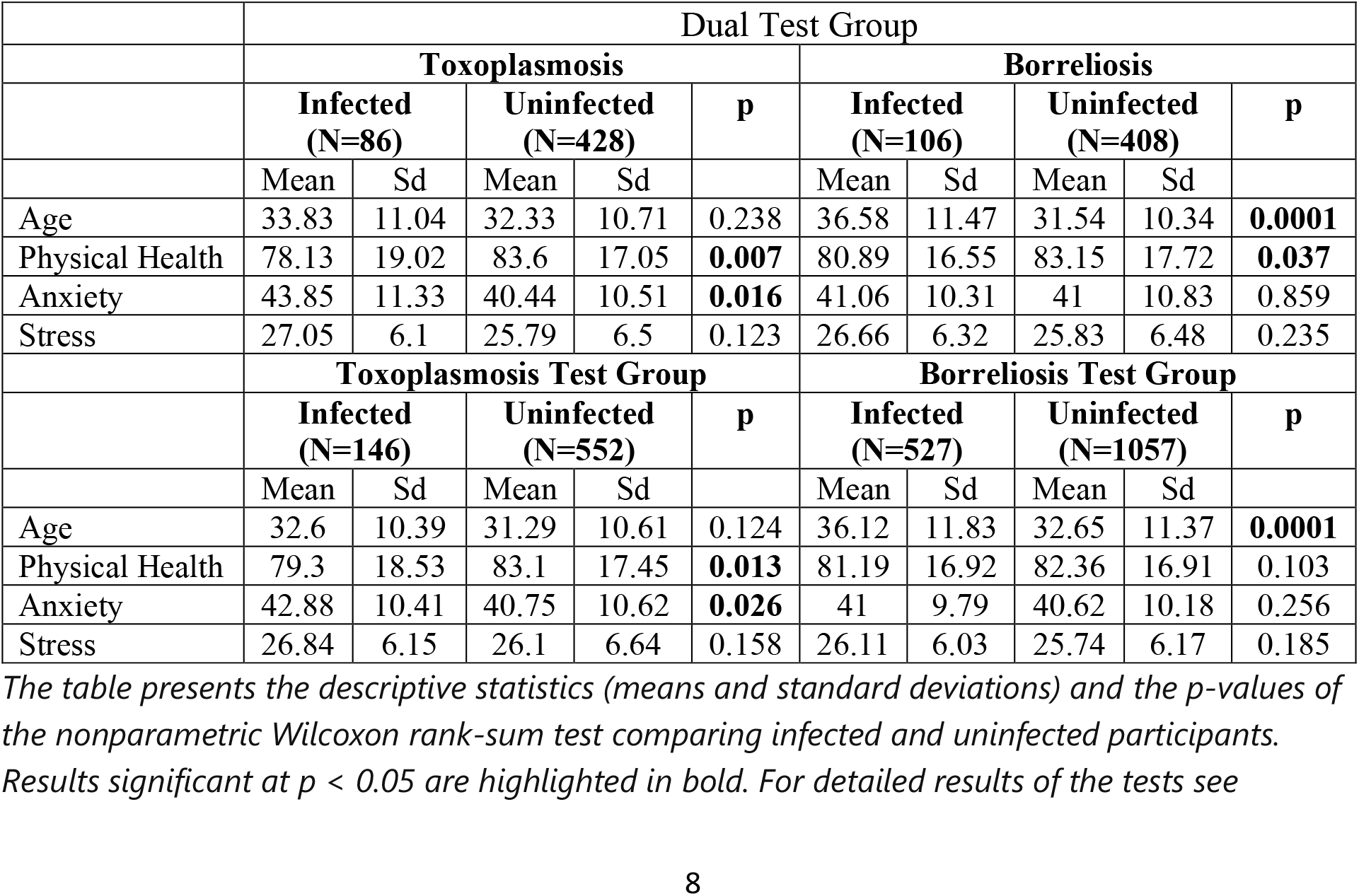
Descriptive Statistics for Toxoplasmosis, Borreliosis, and Dual Test groups

|  | Dual Test Group |  |  |  |  |  |  |  |  |  |
| --- | --- | --- | --- | --- | --- | --- | --- | --- | --- | --- |
|  | Toxoplasmosis |  |  |  |  | Borreliosis |  |  |  |  |
|  | Infected<br>(N=86) |  | Uninfected<br>(N=428) |  | p | Infected<br>(N=106) |  | Uninfected<br>(N=408) |  | p |
|  | Mean | Sd | Mean | Sd |  | Mean | Sd | Mean | Sd |  |
| Age | 33.83 | 11.04 | 32.33 | 10.71 | 0.238 | 36.58 | 11.47 | 31.54 | 10.34 | **0.0001** |
| Physical Health | 78.13 | 19.02 | 83.6 | 17.05 | **0.007** | 80.89 | 16.55 | 83.15 | 17.72 | **0.037** |
| Anxiety | 43.85 | 11.33 | 40.44 | 10.51 | **0.016** | 41.06 | 10.31 | 41 | 10.83 | 0.859 |
| Stress | 27.05 | 6.1 | 25.79 | 6.5 | 0.123 | 26.66 | 6.32 | 25.83 | 6.48 | 0.235 |
|  | Toxoplasmosis Test Group |  |  |  |  | Borreliosis Test Group |  |  |  |  |
|  | Infected<br>(N=146) |  | Uninfected<br>(N=552) |  | p | Infected<br>(N=527) |  | Uninfected<br>(N=1057) |  | p |
|  | Mean | Sd | Mean | Sd |  | Mean | Sd | Mean | Sd |  |
| Age | 32.6 | 10.39 | 31.29 | 10.61 | 0.124 | 36.12 | 11.83 | 32.65 | 11.37 | **0.0001** |
| Physical Health | 79.3 | 18.53 | 83.1 | 17.45 | **0.013** | 81.19 | 16.92 | 82.36 | 16.91 | 0.103 |
| Anxiety | 42.88 | 10.41 | 40.75 | 10.62 | **0.026** | 41 | 9.79 | 40.62 | 10.18 | 0.256 |
| Stress | 26.84 | 6.15 | 26.1 | 6.64 | 0.158 | 26.11 | 6.03 | 25.74 | 6.17 | 0.185 |

The table presents the descriptive statistics (means and standard deviations) and the p-values of the nonparametric Wilcoxon rank-sum test comparing infected and uninfected participants. Results significant at $p < 0.05$ are highlighted in bold. For detailed results of the tests see

Crosstabulation of toxoplasmosis and sex (all men = 172 (24.64%), all women = 526 (75.36%); infected men = 28 (16.27%), infected women = 118 (22.43%)) showed a non-significant trend towards a higher prevalence of toxoplasmosis in women (χ2(1) = 2.967, p = 0.084). The same analysis for borreliosis and sex (all men = 650 (41.04%), all women = 934 (58.96%); infected men = 206 (31.69%), infected women = 321 (34.36%)) yielded no effect of sex on prevalence of borreliosis (χ^2^(1)= 1.236, p = 0.266). In the Dual Test Group, i.e. in participants who reported data on both infections, the prevalence of *Borrelia* infection was higher among individuals infected with *Toxoplasma* (34 subjects (39.53%)) compared to those not infected with *Toxoplasma* (72 subjects (16.82 %)). This disparity was statistically significant (χ^2^_(1)_= 22.566, p<0.0001).

### 3.2. Stress and anxiety in infected and uninfected subjects

#### 3.2.1. Association between toxoplasmosis and borreliosis and anxiety and stress: MANCOVA and ANCOVA test results

The results of Box’s test for equality of covariance matrices and Levene’s test for equality of error variances for stress were satisfactory for the Toxoplasmosis Test Group, Borreliosis Test Group, and the Dual Test Group. The analyses indicated a breach of the assumption of equal error variances for anxiety within the Borreliosis Test group, an issue not encountered in the Toxoplasmosis Test group or the Dual Test group. It is important to note, however, that, notwithstanding this irregularity, the potential for a Type I Error remains minimal, since the effects of borreliosis on stress and anxiety did not reach statistical significance in the Borreliosis Test group, as detailed below. Moreover, the nonparametric test (partial Kendall correlation with age and sex controlled performed separately for toxoplasmosis and borreliosis and men and women) provided practically identical tests as parametric tests (MANCOVA, ANCOVA, and path analysis).

MANCOVA models were constructed for the dependent variables of anxiety and stress, and the independent variables of sex, either toxoplasmosis or borreliosis, or both toxoplasmosis and borreliosis, controlled for age.

The MANCOVA analysis for the Toxoplasmosis Test group revealed significant effects for toxoplasmosis and age; however, the interaction between toxoplasmosis and sex did not prove significant. The MANCOVA modeling analysis for the Borreliosis Test group revealed that neither borreliosis nor its interaction with sex was significant, whereas age and sex themselves had significant effects. MANCOVA results for the Dual Test group demonstrated that the effect of toxoplasmosis was again significant, but the effect of borreliosis, the interactions between sex and toxoplasmosis, sex and borreliosis, toxoplasmosis and borreliosis, and toxoplasmosis, borreliosis, sex, and age proved non-significant. For more details on all MANCOVA models see Table 2.

**Table 2.**
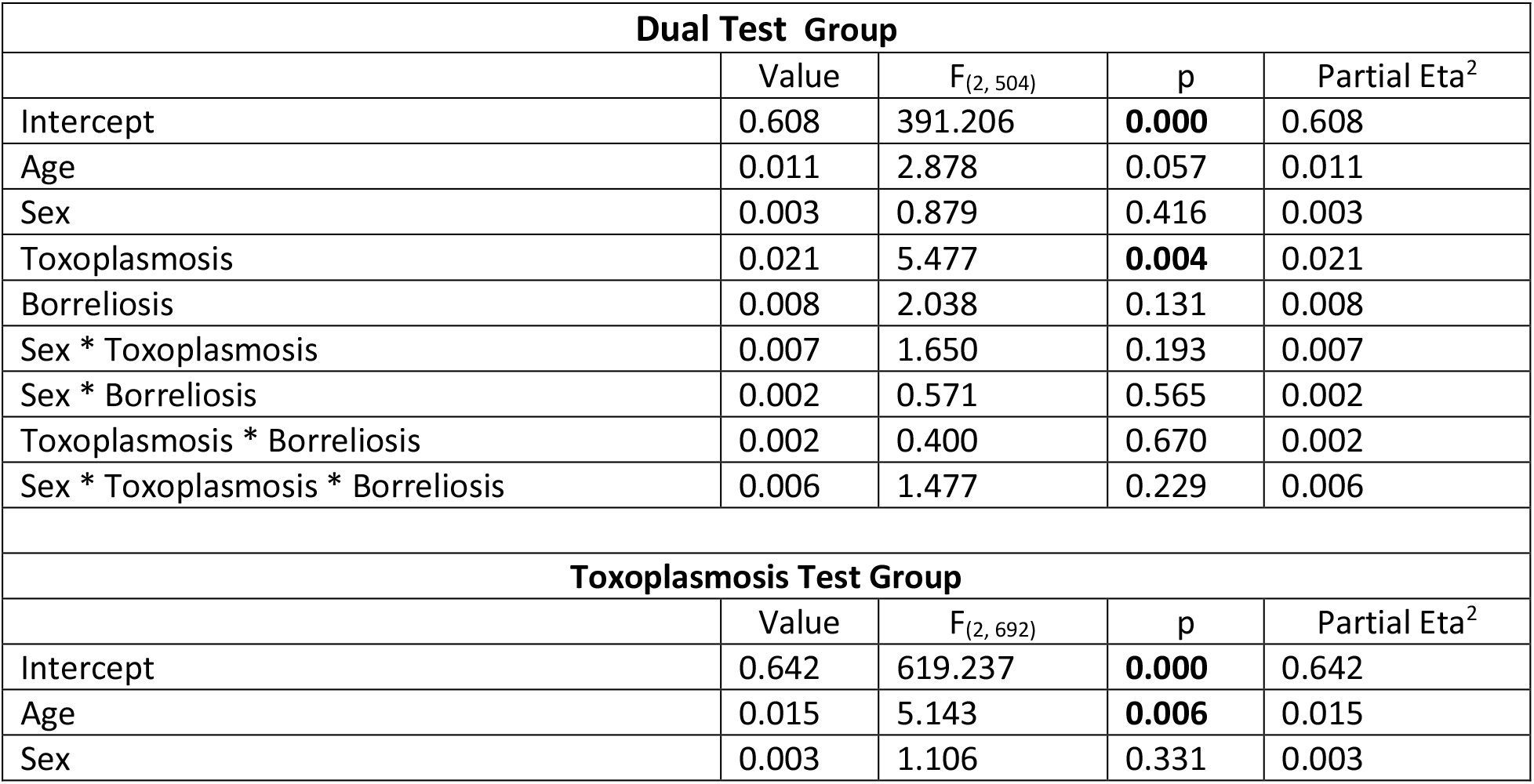
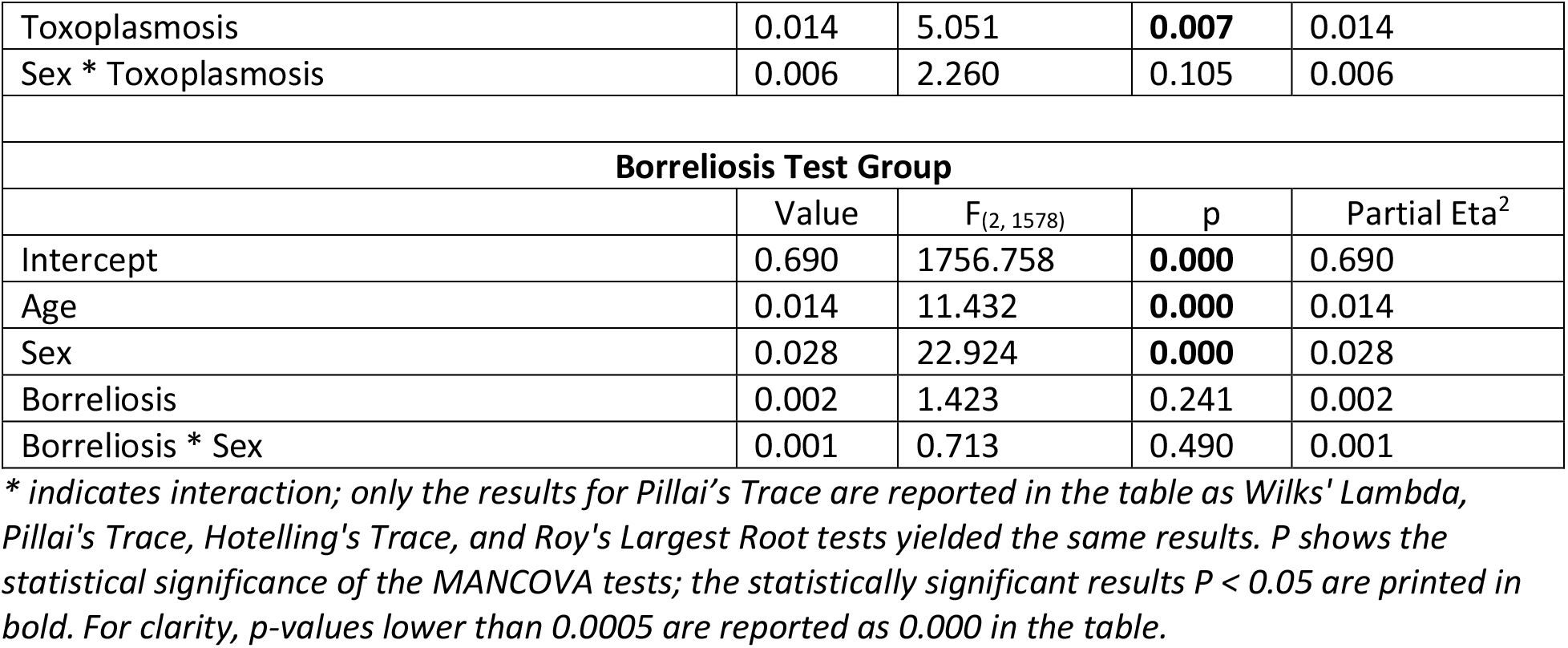
Results of multivariate Analysis of covariance for anxiety and stress

The ANCOVA follow-up univariate analysis for the Toxoplasmosis Test group showed that the effects of toxoplasmosis and its interaction with sex were significant for anxiety, so that higher levels of anxiety were present in infected individuals, and, as the follow-up analysis showed, infected men were the most affected. Although the toxoplasmosis effect also proved significant for stress showing that infected individuals experienced higher levels of stress, its interaction with sex did not. The ANCOVA follow-ups for Borreliosis Test group also revealed that neither borreliosis nor its interaction with sex were significant for anxiety, and the same held for stress. The ANCOVA results for the Dual Test group indicated that the effects of toxoplasmosis, borreliosis, the interaction between toxoplasmosis and borreliosis, the interaction between borreliosis and sex, the interaction between toxoplasmosis and sex, and the interaction among borreliosis, toxoplasmosis, and sex were not significant for either anxiety or stress ANCOVA tests also showed that older participants expressed significantly lower anxiety and stress. For the strength, significance, and direction of the effects see Table 3 and Fig. 2.

**Fig. 2.**
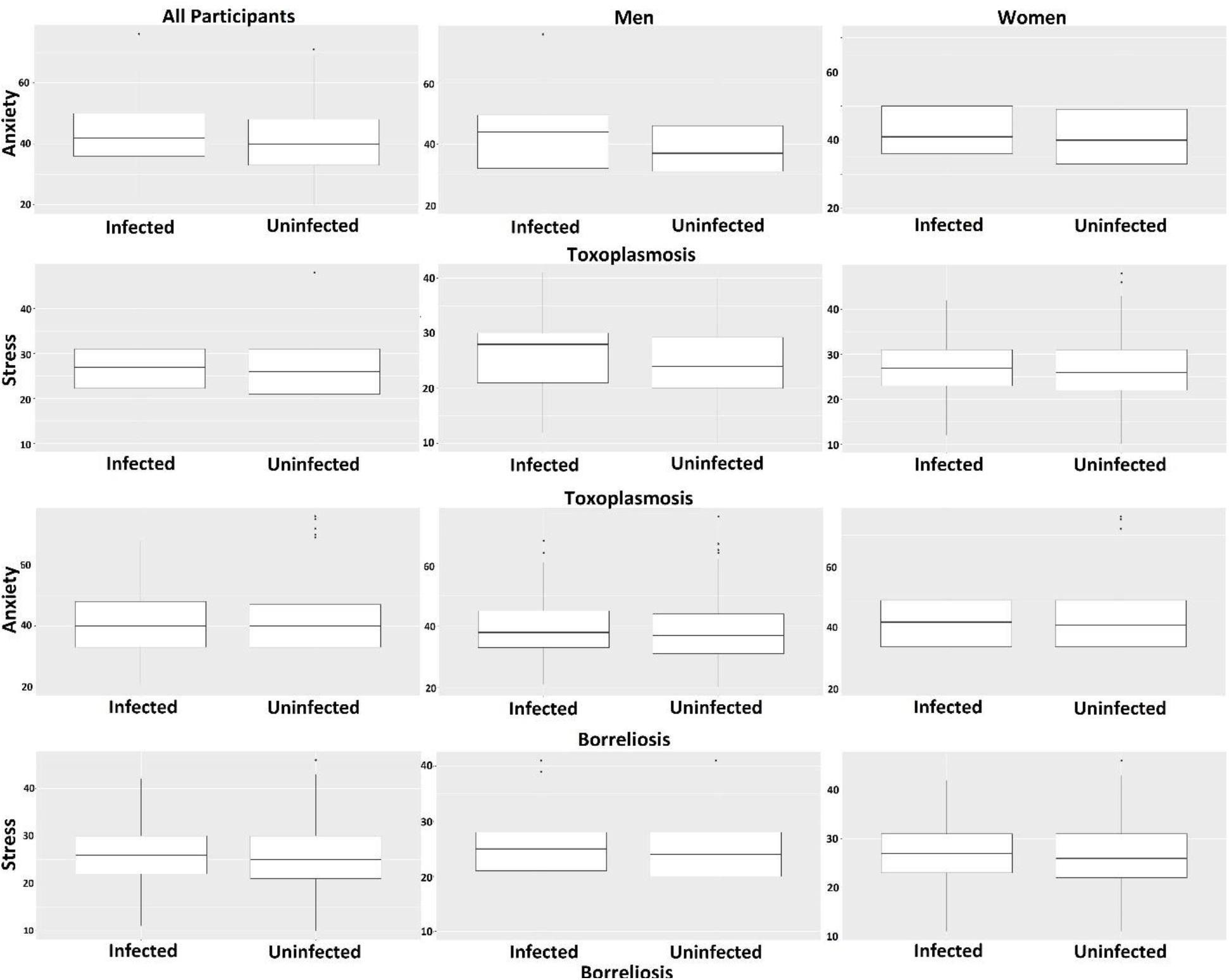
Effects of *Toxoplasma* and *Borrelia* infections on anxiety and stress

**Table 3.**
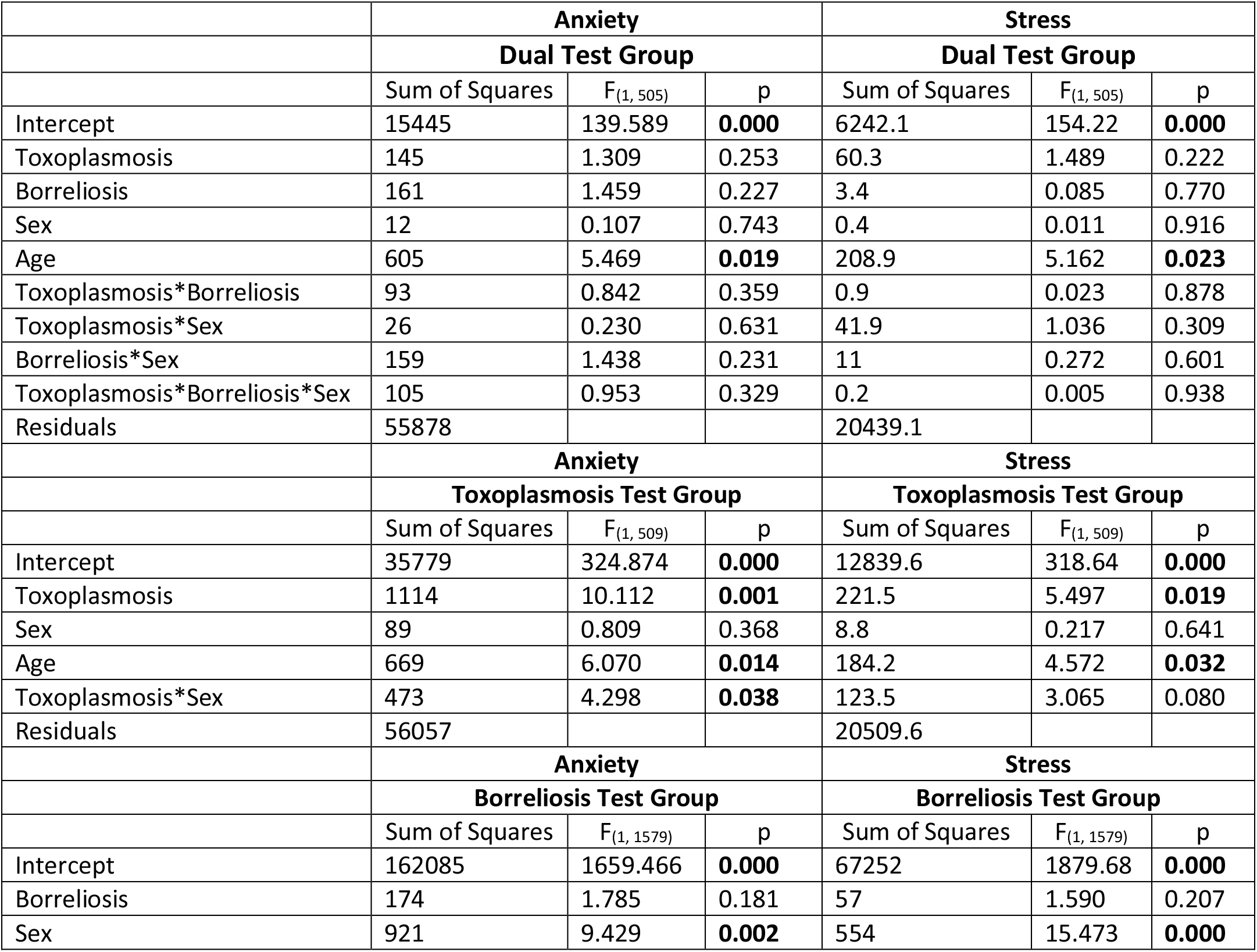
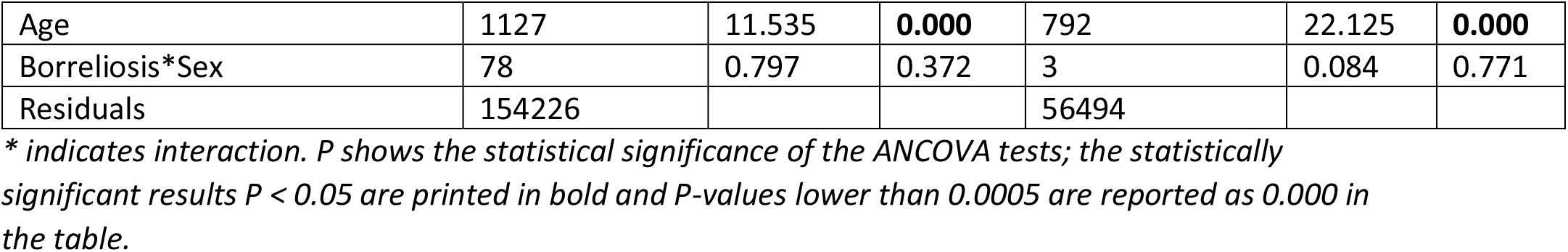
Results of univariate analysis of covariance for anxiety and stress *Boxplots show the distributions of the levels of anxiety (rows 1 and 3) and stress (rows 2 and 4) for the infected and uninfected subjects (split by three categories of ‘all participants’, ‘men’, and ‘women’ in columns 1, 2, and 3, respectively) in the Toxoplasmosis Test group (rows 1 and 2) and also in the Borreliosis Test group (rows 3 and 4). Boxplots show the 25^th^ and 75^th^ percentiles as box boundaries, with a line marking the median inside each box. The whiskers show the range of variability of the distributions within 95% confidence intervals*.

|  | **Anxiety** |  |  | **Stress** |  |  |
| --- | --- | --- | --- | --- | --- | --- |
|  | **Dual Test Group** |  |  | **Dual Test Group** |  |  |
| | Sum of Squares | $F_{(1, 505)}$ | p | Sum of Squares | $F_{(1, 505)}$ | p |
| Intercept | 15445 | 139.589 | **0.000** | 6242.1 | 154.22 | **0.000** |
| Toxoplasmosis | 145 | 1.309 | 0.253 | 60.3 | 1.489 | 0.222 |
| Borreliosis | 161 | 1.459 | 0.227 | 3.4 | 0.085 | 0.770 |
| Sex | 12 | 0.107 | 0.743 | 0.4 | 0.011 | 0.916 |
| Age | 605 | 5.469 | **0.019** | 208.9 | 5.162 | **0.023** |
| Toxoplasmosis*Borreliosis | 93 | 0.842 | 0.359 | 0.9 | 0.023 | 0.878 |
| Toxoplasmosis*Sex | 26 | 0.230 | 0.631 | 41.9 | 1.036 | 0.309 |
| Borreliosis*Sex | 159 | 1.438 | 0.231 | 11 | 0.272 | 0.601 |
| Toxoplasmosis*Borreliosis*Sex | 105 | 0.953 | 0.329 | 0.2 | 0.005 | 0.938 |
| Residuals | 55878 |  |  | 20439.1 |  |  |
|  | **Anxiety** |  |  | **Stress** |  |  |
|  | **Toxoplasmosis Test Group** |  |  | **Toxoplasmosis Test Group** |  |  |
| | Sum of Squares | $F_{(1, 509)}$ | p | Sum of Squares | $F_{(1, 509)}$ | p |
| Intercept | 35779 | 324.874 | **0.000** | 12839.6 | 318.64 | **0.000** |
| Toxoplasmosis | 1114 | 10.112 | **0.001** | 221.5 | 5.497 | **0.019** |
| Sex | 89 | 0.809 | 0.368 | 8.8 | 0.217 | 0.641 |
| Age | 669 | 6.070 | **0.014** | 184.2 | 4.572 | **0.032** |
| Toxoplasmosis*Sex | 473 | 4.298 | **0.038** | 123.5 | 3.065 | 0.080 |
| Residuals | 56057 |  |  | 20509.6 |  |  |
|  | **Anxiety** |  |  | **Stress** |  |  |
|  | **Borreliosis Test Group** |  |  | **Borreliosis Test Group** |  |  |
| | Sum of Squares | $F_{(1, 1579)}$ | p | Sum of Squares | $F_{(1, 1579)}$ | p |
| Intercept | 162085 | 1659.466 | **0.000** | 67252 | 1879.68 | **0.000** |
| Borreliosis | 174 | 1.785 | 0.181 | 57 | 1.590 | 0.207 |
| Sex | 921 | 9.429 | **0.002** | 554 | 15.473 | **0.000** |
| Age | 1127 | 11.535 | **0.000** | 792 | 22.125 | **0.000** |
| Borreliosis*Sex | 78 | 0.797 | 0.372 | 3 | 0.084 | 0.771 |
| Residuals | 154226 |  |  | 56494 |  |  |

*\* indicates interaction. P shows the statistical significance of the ANCOVA tests; the statistically significant results $P < 0.05$ are printed in bold and P-values lower than 0.0005 are reported as 0.000 in the table.*

#### 3.2.2. Could impaired physical health mediate the effects of infections on stress and anxiety? Partial Kendall correlation results

The Stress-Coping Hypothesis suggests a mediating role of physical health and stress in the effects of toxoplasmosis on behavioral variables and personality traits. Concurrently, the data presented in Table 1 indicate that physical health is indeed influenced by toxoplasmosis. To assess this hypothesis and at the same time to confirm the results of parametric tests with more robust (assumption-free) nonparametric tests, we analyzed the effects of infections on stress and anxiety using partial Kendall correlation test. To reveal the possible mediating effect of physical health impairment, we juxtaposed the outcomes of analyses that controlled solely for age and sex against those that also controlled for physical health (Table 4).

**Table 4.** Partial Kendall Tau correlations and their attributed p-values *Positive Kendall Tau partial correlation coefficients indicate a rise in anxiety or stress levels in association with toxoplasmosis or borreliosis infection. Conversely, negative coefficients signify a reduction in anxiety or stress levels correlated with these infections. Significant p values are highlighted in bold*

| Dual Test Group |  |  |  |  |  |  |  |  |  |  |  |  |  |
| --- | --- | --- | --- | --- | --- | --- | --- | --- | --- | --- | --- | --- | --- |
|  |  | Covariates Age and Sex |  |  |  |  |  | Covariates physical health, Age and Sex |  |  |  |  |  |
|  |  | All |  | Women |  | Men |  | All |  | Women |  | Men |  |
|  |  | Anxiety | Stress | Anxiety | Stress | Anxiety | Stress | Anxiety | Stress | Anxiety | Stress | Anxiety | Stress |
| Toxoplasmosis | $\tau$ | 0.086 | 0.054 | 0.053 | 0.025 | 0.171 | 0.132 | 0.058 | 0.029 | 0.032 | 0.007 | 0.122 | 0.088 |
|  | p | **0.003** | 0.063 | 0.129 | 0.464 | **0.002** | **0.016** | **0.048** | 0.314 | 0.349 | 0.833 | **0.027** | 0.111 |
| Borreliosis | $\tau$ | 0.006 | 0.044 | 0.006 | 0.05 | 0.008 | 0.022 | -0.015 | 0.027 | -0.021 | 0.029 | 0.003 | 0.018 |
|  | p | 0.814 | 0.134 | 0.845 | 0.152 | 0.879 | 0.681 | 0.602 | 0.356 | 0.547 | 0.407 | 0.956 | 0.736 |
| Toxoplasmosis Test Group |  |  |  |  |  |  |  |  |  |  |  |  |  |
|  |  | Covariates Age and Sex |  |  |  |  |  | Covariates physical health, Age and Sex |  |  |  |  |  |
|  |  | All |  | Women |  | Men |  | All |  | Women |  | Men |  |
|  |  | Anxiety | Stress | Anxiety | Stress | Anxiety | Stress | Anxiety | Stress | Anxiety | Stress | Anxiety | Stress |
| Toxoplasmosis | $\tau$ | 0.068 | 0.042 | 0.044 | 0.02 | 0.141 | 0.114 | 0.044 | 0.021 | 0.024 | 0.002 | 0.106 | 0.084 |
|  | p | **0.007** | 0.095 | 0.123 | 0.478 | **0.006** | **0.026** | 0.079 | 0.404 | 0.407 | 0.941 | **0.039** | 0.102 |
| Borreliosis Test Group |  |  |  |  |  |  |  |  |  |  |  |  |  |
|  |  | Covariates Age and Sex |  |  |  |  |  | Covariates physical health, Age and Sex |  |  |  |  |  |
|  |  | Anxiety | Stress | Anxiety | Stress | Anxiety | Stress | Anxiety | Stress | Anxiety | Stress | Anxiety | Stress |
| Borreliosis | $\tau$ | 0.026 | 0.034 | 0.013 | 0.023 | 0.051 | 0.047 | 0.02 | 0.03 | 0.011 | 0.021 | 0.042 | 0.04 |
|  | p | 0.118 | **0.041** | 0.538 | 0.285 | 0.051 | 0.068 | 0.211 | 0.073 | 0.605 | 0.316 | 0.109 | 0.121 |

The initial, less complex model without control for physical health for the Toxoplasmosis Test group demonstrated that toxoplasmosis significantly impacts anxiety. The associations between toxoplasmosis and both stress and anxiety were stronger and statistically significant in men, while in women, they were weaker and did not reach significance. When the model was expanded to include the covariates of age, sex, and physical health, the observed effects of toxoplasmosis diminished. Only the impact of the infection on anxiety in men remained significant. Similar analyses for borreliosis did not identify any significant effect except for its correlation with stress across the entire sample, including both men and women. Again, the significant correlation and also trends observed were significantly attenuated when physical health was controlled (see Table 4).

#### 3.2.3. Direct and indirect effects of infections on health, stress, and anxiety - Path Analysis

The outcomes of the partial Kendall analysis indicate that physical health plays a pivotal role in mediating the correlations of infections with stress and anxiety. However, this analysis neither elucidates the mechanisms behind such mediation nor excludes the possibility that the infections may also have direct effects on stress or its significant psychological symptom, anxiety. To further investigate these critical issues, we employed structural modeling, specifically path analysis. The impacts of toxoplasmosis and borreliosis were analyzed separately. The models included variables such as age, physical health, anxiety, stress, and infection status (toxoplasmosis or borreliosis). In additional post hoc analyses, the effect of sex was evaluated by conducting identical analyses for each sex separately. The findings depicted in Figures 3 and 4 suggest that toxoplasmosis exerts only indirect effects on stress, these being mediated through its direct impacts on physical health. The effect of toxoplasmosis on anxiety also seems to be mostly indirect, mediated through physical health. However, a substantial portion of the effect of toxoplasmosis on anxiety can be ascribed to its physical health-mediated influence on stress and the pronounced impact of stress on anxiety. Post hoc analyses indicate that this pattern slightly differs between men (Fig. 3a) and women (Fig. 3b). In women, the physical health-mediated effect of toxoplasmosis on stress is nonsignificant and much weaker than in men, and its direct effect on stress is even weaker (and negative instead of positive).

**Figure 3.**
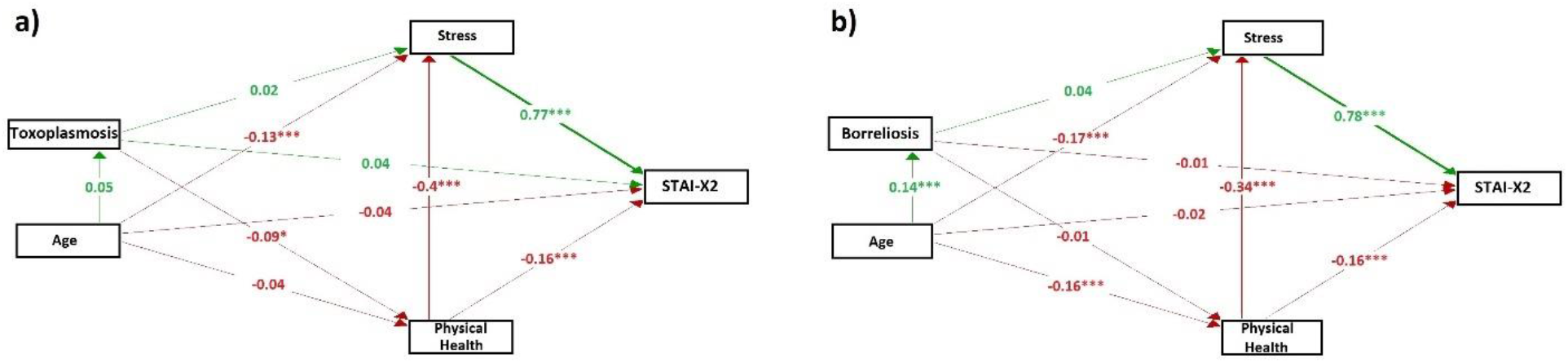
Path analysis for Toxoplasmosis Test and Borreliosis Test Groups *Positive path coefficients (green arrows) indicate that an increase in the source variable leads to an increase in the dependent variable. Conversely, negative coefficients (red arrows) imply that an increase in the source variable results in a decrease in the dependent variable. For dichotomous variables like toxoplasmosis and borreliosis, positive coefficients indicate that infection is associated with an increase in the dependent variable, while negative coefficients indicate that infection is associated with a decrease in the dependent variable. * indicates p<0.05; ** indicates p<0.01; *** indicates p<0.001*

**Figure 4.**
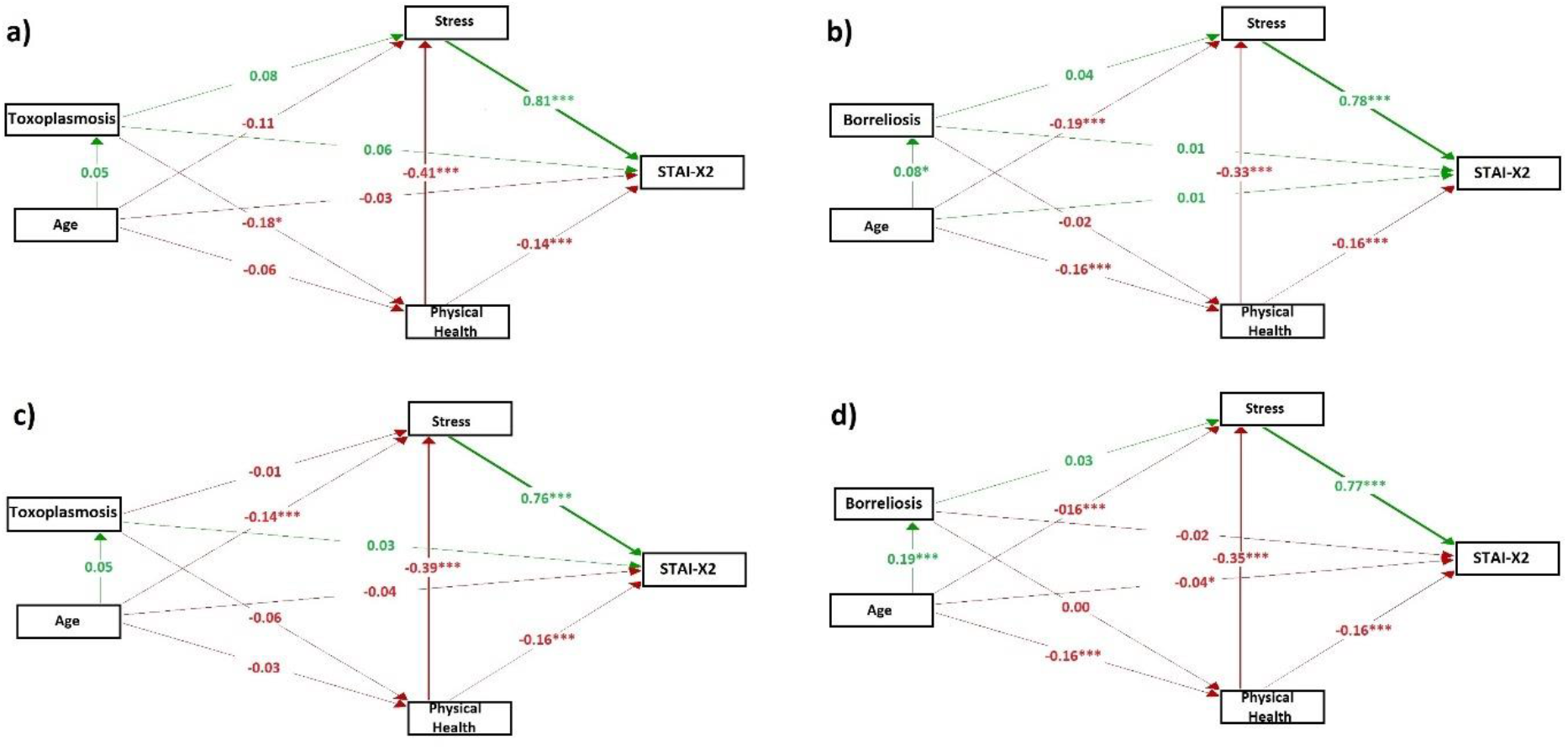
Path analysis for men and women in Dual Test Groups *Parts a and b depict models for men, whereas parts c and d present models for women. For additional explanation, refer to the legend for Fig. 2*.

In contrast, borreliosis exerts a significant direct effect on stress, and a weak nonsignificant effect on physical health. Here, the overarching trend observed is strikingly similar in men (Fig. 3c) and women (Fig. 3d). For a more detailed interpretation of the results of path analyses see Discussion.

## 4. Discussion

### 4.1. Association of *Toxoplasma* infection with higher stress and anxiety

Using standard psychological questionnaires; namely, the STAI-X2 and PSS, we measured perceived stress and anxiety in 1,768 men and women who had previously undergone laboratory testing for toxoplasmosis or borreliosis and provided information about the results of these tests. The collected data were used in our study to test the Stress-Coping Hypothesis (Lindová et al., 2010; Lindová et al., 2006). This hypothesis suggests that many psychological differences between individuals infected with the *Toxoplasma* parasite, particularly those that are directionally opposite in men and women, are not the result of the parasite’s manipulative activity aimed at facilitating transmission from the infected to the uninfected host, but are induced by chronic stress caused by the long-term impact of the infection on physical health. The outputs of both parametric and non-parametric tests were consistent with the Stress-Coping Hypothesis (Lindová et al., 2010; Lindová et al., 2006). While the direct effect of infection on stress and, consequently, on its primary psychological manifestation, anxiety, was relatively weak, toxoplasmosis had a strong effect on physical health. Deterioration in physical health had a direct effect on stress, resulting in higher levels of stress (and its manifestation, anxiety) in *Toxoplasma*-infected individuals. In our sample, both direct and indirect effects of toxoplasmosis were stronger in men than in women.

### 4.2. Association of *Borrelia* infection with higher stress and anxiety

Although the public is more concerned about the long-term health consequences of borreliosis than toxoplasmosis, the association of borreliosis with physical health was minimal (non-existent in women) and the observed weaker positive association between infection and anxiety in men was likely due to a higher incidence of toxoplasmosis in individuals infected with borreliosis, as discussed below. This confirms that the impact of toxoplasmosis is likely specific and, from a public health perspective, more significant than the influence of other similarly widespread chronic infections. This conclusion is in line with the results of previously published studies. These have shown that differences in the prevalence of toxoplasmosis explain 23% of the variance in overall disease burden in different European countries (Flegr et al., 2014) and that the prevalence of toxoplasmosis is the third most important predictor of the ratio of male to female births (with a lower ratio serving as a proxy for impaired health) among the 15 factors studied (Dama, Novakova, & Flegr, 2016).

The absence of associations between infection and health in the case of borreliosis calls into question the hypothesis that the connection between infection and health is simply an artifact of questionnaire methodologies. It specifically casts doubt on the notion that individuals with heightened health awareness are predisposed to report worse health and the presence of an infection, thereby supporting the likelihood that, on average, *Toxoplasma*-infected individuals truly experience poorer health and, secondarily, higher stress compared to those uninfected.

### 4.3. Why the association of toxoplasmosis with anxiety seems stronger than with stress?

The overall (direct + indirect) association of toxoplasmosis with stress was consistently weaker than with anxiety. This seemingly contradicts our hypothesis, which posits that the association with stress is primary and that with anxiety is secondary, mediated by the impact of stress on anxiety (Hussenoeder et al., 2022). In reality, the strength of a statistical association—reflected in measures such as partial Eta squared values, absolute Kendall Tau values, or absolute path coefficients—is influenced not only by the true strength of the association but also by the precision with which the studied variables are measured. In our study, the State-Trait Anxiety

Inventory-Revised X2, which consists of 20 questions, demonstrated higher reliability (Omega = 0.94) compared to the Perceived Stress Scale (PSS) that contains only 10 questions (Omega = 0.90). This finding suggests that the STAI-X2 may more accurately estimate psychological states than the PSS. Consequently, the measured strength of the association of infection with stress could appear lower than with anxiety in our analysis, even if the actual strength of the association with stress is higher.

### 4.4. Why effects of toxoplasmosis were stronger in men than in women?

The strength of the association between toxoplasmosis and other variables was significantly stronger in men than in women. This aligns with the results of most previous studies (Flegr & Horáček, 2020; Flegr & Hrdý, 1994; Flegr, Šebánková, Příplatová, Chvátalová, & Kaňková, 2018; Lindová, Příplatová, & Flegr, 2012; Příplatová, Šebanková, & Flegr, 2014). In the case of the association of latent toxoplasmosis with morphological and some behavioral traits, this could be due to the physiological mechanism of some behavioral effects of toxoplasmosis.

Experiments with laboratory-infected rats have demonstrated that *Toxoplasma* infection often increases testosterone synthesis in the testes, leading to elevated testosterone levels which are directly or indirectly responsible for the observed behavioral changes. In female rats and castrated male rats, such an increase in testosterone levels and the corresponding behavioral changes do not occur (Lim, Kumar, Dass, & Vyas, 2013). Increased testosterone levels have also been observed in *Toxoplasma*-infected men, whereas infected women exhibited lower testosterone levels (Flegr, Lindová, & Kodym, 2008). The higher stress and greater intensity of stress-dependent psychological changes in men could be related to an increased testosterone level. High levels of testosterone could lead to immunosuppression and therefore more frequent illnesses, which could deteriorate physical health, increase stress levels, and secondarily increase anxiety. The health deterioration observed, although to a lesser extent, in infected women suggests that *Toxoplasma* impacts physical health and thereby stress also through mechanisms beyond just elevated testosterone synthesis in the testes.

Of course, it is possible that the greater intensity of *Toxoplasma*-associated manifestations in men compared to women could have entirely different causes. Most women learned of their *Toxoplasma* positivity during preventive screening in pregnancy, whereas men were tested for toxoplasmosis while investigating possible causes of health issues. For example, among 2,044 participants of a similar internet study, 37.6% women were screened for toxoplasmosis in relation to their pregnancy, 40.3% men and 34.3% women were tested in our lab during participation in various research projects, and 49.4% men and 20.7% women were tested in relation to their health problems (Flegr & Preiss, 2019). Therefore, it is possible that, on average, male participants suffered from a clinically more severe form of toxoplasmosis than female participants.

It can be also hypothesized that due to different stress-coping strategies in men and women, the questionnaires may measure stress and anxiety more accurately in men than in women. When testing the reliability of each questionnaire separately for men and women, we found the results to be nearly identical across genders, although the reliability differed between the two questionnaires, see the Materials and Methods section. However, it’s important to acknowledge that a questionnaire’s statistical reliability doesn’t automatically guarantee its construct validity. This means a questionnaire intended to measure anxiety or stress might not do so with equal validity for both men and women. This discrepancy arises because men and women typically encounter different stressors, utilize distinct coping strategies, are socialized into different narratives around anxiety and stress (McLean & Anderson, 2009), and may interpret the same questionnaire item differently.

### 4.5. Do some symptoms of senescence actually reflect the increasing prevalence of toxoplasmosis in age strata?

Path analysis revealed that age, unsurprisingly, correlates positively with the likelihood of having been diagnosed with borreliosis and toxoplasmosis during one’s lifetime. However, a potentially significant finding emerged from comparing the correlation of age with physical health in the path analysis model including toxoplasmosis and the model including borreliosis. In the model concerning borreliosis—a condition which had no effect on physical health—a strong negative impact of age on physical health was observed. In the model incorporating toxoplasmosis instead of borreliosis—where toxoplasmosis effects were explicitly controlled— the impact of age on physical health was observed to be four times weaker. This suggests that the influence of age on physical health, commonly seen as aging manifestations, might be significantly impacted by toxoplasmosis. With the increasing probability of contracting toxoplasmosis with age, and its latent manifestations often being cumulative (Flegr, Hrdá, & Kodym, 2005; Flegr & Hrdý, 1994; Flegr et al., 1996; Lindová et al., 2012), it is conceivable that some health deteriorations commonly attributed to aging could actually result from the pathological effects of toxoplasmosis.

The public health impact of toxoplasmosis on the whole population could be substantial. On average, about one-third of the population is infected with toxoplasmosis. Given that its prevalence increases with age, it is likely that more than half of the population in older age groups are infected. Moreover, in older infected humans, the level of anamnestic anti- *Toxoplasma* IgG antibodies decreases in some individuals below the positivity threshold. This is manifested, for example, by the fact that although the seroprevalence of toxoplasmosis should increase with age, it only rises up to a certain age and then begins to decrease in the respective age groups (Flegr, 2017; Kolbeková, Kourbatova, Novotná, Kodym, & Flegr, 2007). This indicates that the prevalence values commonly reported, particularly for the oldest age groups, are much lower than the actual prevalence of infected individuals within those age categories.

### 4.6. Are our findings regarding significantly worse health of individuals with latent toxoplasmosis consistent with current knowledge?

Independent partial clinical studies and extensive systematic research, primarily conducted in the last decade, indeed show that the prevalence of latent toxoplasmosis significantly impacts public health; for a review, see (Flegr et al., 2014). For example, a comprehensive systematic cross-sectional online study, which examined the incidence of all disease types, reveals that a significant portion of diseases occur more frequently in individuals diagnosed with latent toxoplasmosis compared to those without the infection. Specifically, the study showed that, of the 134 disorders reported by at least 10 participants, 77 were significantly more common in *Toxoplasma*-infected participants than in *Toxoplasma*-free participants. The study also revealed that 333 infected subjects scored significantly worse than 1,153 uninfected controls in 28 of 29 health-related variables (Flegr & Escudero, 2016). Similar results were provided by a large ecological study that examined the correlation of the prevalence of toxoplasmosis in 88 countries with health impacts, specifically age-standardized Disability Adjusted Life Year (DALY), for 128 diseases monitored by WHO over the long term (Flegr et al., 2014). The study showed that 23 of these diseases statistically significantly correlate with the prevalence of toxoplasmosis in individual countries. Such diseases include major public health concerns such as inflammatory heart disorders, ischemic heart disease, cerebrovascular disease, prostate cancer, and epilepsy. The study also showed that differences in the occurrence of toxoplasmosis in individual countries, after filtering out confounding variables such as Gross Domestic Product per capita (GDP), geolatitude, and humidity, explain 23% of all variability in overall morbidity in the 29 European countries included in the study.

### 4.7. Possible reasons for a positive association between *Toxoplasma* and *Borrelia* infection

An incidental finding of the current study was the demonstration of a strong correlation between *Toxoplasma* and *Borrelia* infections; individuals previously diagnosed with one parasite had a significantly higher probability of being diagnosed with the other. This positive association may explain the seemingly direct positive association of borreliosis with stress (path coefficient = 0.04, p = 0.032). Most likely, a positive test for borreliosis simply indicated a higher likelihood of toxoplasmosis positivity. Toxoplasmosis was not included in the model, and moreover, the majority of study participants were not tested for toxoplasmosis. Although the analysis might suggest that borreliosis positivity is associated with worsened health, it was actually toxoplasmosis, not borreliosis, that impacted physical health and subsequently increased stress.

A positive association between *Toxoplasma* and *Borrelia* seropositivity was also reported in previous studies (Flegr & Horáček, 2018; Flegr et al., 2023). The existence of this association can be explained in at least three fundamentally different ways. The first possibility is that infection by one of the pathogens could increase the likelihood of infection by the other pathogen, most likely through the induction of changes in the host’s immune system functioning – immunomodulation leading to reduced responsiveness or non-responsiveness to certain types of antigens. Indeed, numerous studies have shown that toxoplasmosis induces dramatic changes in the immune system functioning of both infected animals and humans (Flegr & Stříž, 2011; Š. Kaňková, Holáň, Zajícová, Kodym, & Flegr, 2010; Tomita et al., 2021).

The second possibility is that both pathogens could be transmitted to humans simultaneously – for instance, through tick bites. Borreliosis in the Czech Republic is transmitted by *Ixodes ricinus* ticks. While this mode of transmission is not yet assumed for toxoplasmosis, *Toxoplasma* DNA has been identified in *Ixodes ricinus* at a rate almost identical to *Borrelia* (12.6% versus 12.7%) (Sroka, Szymanska, & Wojcik-Fatla, 2009). Moreover, both pathogens were detected together in 2.3% of all ticks and 3.8% of adult female ticks. Previous research has also confirmed the transmission of *Toxoplasma* by three tick species: *Dermacentor variabilis*, *D. andersoni*, and *Amblyomma americanum* (Woke, Jacobs, Jones, & Melton, 1953).

The third possibility is that the association could be an artifact of questionnaire studies – a portion of individuals might be more likely to respond positively to both questions, even though they are not actually infected with both pathogens. We attempted to mitigate this risk in the questionnaire. The participants were asked to report only laboratory test results regarding their infection status with the specified pathogens, and the questionnaire was preset to indicate as a default the response ‘I do not know, I am not sure, I have not been tested’. Even so, it is possible that individuals with certain psychological dispositions, such as heightened concerns about their health, perhaps akin to hypochondria, may have tended to answer both questions positively. Therefore, our potentially significant findings should be considered only preliminary until the existence of an association between borreliosis and toxoplasmosis is confirmed through a study in which both infections are diagnosed serologically during the study itself.

### 4.8. Strengths and limitations of the study

The main strength of this study is the large number of participants and the fact that they were not pre-informed that the questionnaire would also contain questions on toxoplasmosis and borreliosis. The entire study, conducted during the first year of the COVID-19 pandemic, was primarily focused on the impact of COVID-19 and pandemic measures on physical health and wellbeing, with the two questions regarding infections by *Toxoplasma* and *Borrelia* placed towards the end of the questionnaire. Thus, participants could not consciously or subconsciously bias the results based on their beliefs about the influence of toxoplasmosis on their physical health.

A significant limitation of the study is the self-selection of participants, which means our sample does not represent a random cross-section of the general population. Consequently, the extent to which the findings can be generalized remains uncertain. However, it is important to emphasize that the self-selection of participants was random with respect to *Toxoplasma* and *Borrelia* infection, as participants were not aware that the questionnaire would include questions on these variables.

Another limitation is that the anonymous study participants self-reported their laboratory test results for toxoplasmosis and borreliosis, and this information could not be verified. At least in the case of borreliosis, such self-reported information is more accurate than if participants had been tested in the study, as levels of anti-*Borrelia* antibodies often disappear or fluctuate significantly over time after infection. To what extent the same applies to toxoplasmosis is not clear, but reversion to seronegative status is occasionally reported in the literature. However, our results, specifically the decreasing seroprevalence of toxoplasmosis in middle and old age groups, suggest that such reversions might be relatively frequent (Flegr, 2017). It is practically certain that some study participants provided incorrect or outdated data about their infection status; however, the representation of these individuals in the samples is likely very low, according to the findings of one of our earlier studies. In this online study, 1,865 respondents had the option to provide their code, under which they had participated in our studies in the past. This option was utilized by 393 individuals. Verification showed that in 99.2% of cases, the *Toxoplasma* status reported in the questionnaire matched the *Toxoplasma* status determined by our serological tests (Flegr, 2017). Another study demonstrated that the effects of toxoplasmosis on physical and mental health detected in the entire sample of 6,397 individuals were practically identical in strength and direction to those detected in a subsample of 800 individuals who provided their names at the end of the questionnaire, allowing their *Toxoplasma* status to be verified (Flegr & Horáček, 2020).

An additional limitation is that the study was conducted during a pandemic, a time likely marked by increased levels of stress and anxiety within the population. At the beginning of the study and several times throughout the questionnaire, participants were specifically asked to assess their physical health during the ongoing pandemic. For this reason, it is not possible to compare the results obtained during this study with those from normal circumstances, making the extent to which the findings are generalizable uncertain. Furthermore, it is conceivable that our findings do not indicate increased stress and anxiety in *Toxoplasma*-infected subjects but rather imply a diminished capacity in *Toxoplasma*-positive individuals to cope with stressful situations, like the COVID-19 pandemic. It is even possible that the infected individuals objectively face a more severe progression of COVID-19 infection compared to individuals without *Toxoplasma* infection (Flegr, 2021).

Our objective was to assess the impact of infections on long-term psychological states rather than immediate feelings. Hence, we utilized the STAI-X2 variant, which measures anxiety as a trait. However, our questionnaire also included the STAI-6, a brief 6-item inventory that assesses immediate feelings (state) (Marteau & Bekker, 1992). To verify the robustness of our results, we performed analyses with the STAI-6 as an alternative to the STAI-X2; the outcomes were nearly identical (refer to Supplementary Fig. S1). This consistency implies that, within the current study’s context, whether anxiety is measured as a trait or a state appears not to be critically important.

In addition to reporting the results of testing our main hypothesis (higher stress levels in *Toxoplasma*-infected individuals), we also present the findings from additional exploratory analyses. These analyses were not preregistered and may be affected by multiple testing, thus their results should be considered preliminary and require confirmation in independent datasets.

## 5. Conclusion

The Stress-Coping Hypothesis published nearly 20 years ago has so far only been tested and confirmed for one of its two predictions: the deteriorated physical health of individuals with latent toxoplasmosis. Despite an extensive literature review, no published study was found testing its second prediction, increased stress levels in infected individuals. Our study addresses this gap, showing that infected individuals exhibit significantly higher stress and anxiety levels than uninfected ones. The study also reveals that the impact of toxoplasmosis on anxiety and stress is largely mediated by deteriorating physical health in infected individuals. Comparisons of results for toxoplasmosis and borreliosis further indicate that the association between infection, health, and consequently stress is specific to toxoplasmosis and not observed in borreliosis. Furthermore, these comparisons indicate that numerous changes in physical condition commonly ascribed to aging could, in fact, stem from the rising prevalence of toxoplasmosis with age and the accumulative impacts of latent toxoplasmosis.

Our current study strengthens the conclusions of recent research, emphasizing the significant public health impact of latent toxoplasmosis in comparison to other latent infections. Today, around a third of humanity is infected with *Toxoplasma*, and the majority in both developing and developed countries will be infected over their lifetimes. There is no effective drug available for lifelong latent toxoplasmosis in human medicine. However, since *Toxoplasma* can only sexually reproduce in felines, it could be entirely eradicated from the human population using oral vaccines administered to domestic and feral cats. The findings of studies, including ours, suggest that developing such vaccines should be a priority.

## Data Availability

All data are available at figshare https://doi.org/10.6084/m9.figshare.25188788.v1.

## Supplementary materials

**Supplementary figure S1.**
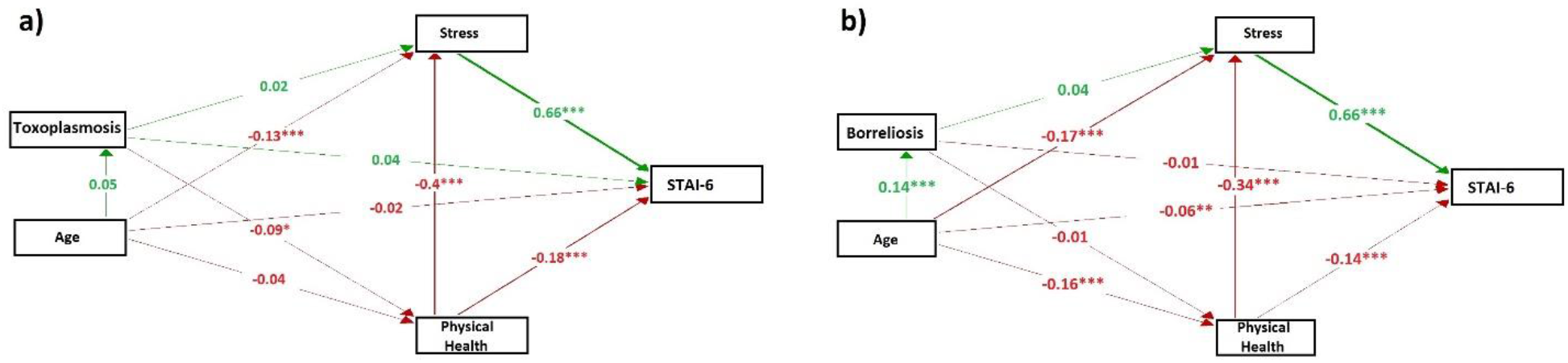
Path analysis for Toxoplasmosis Test and Borreliosis Test Groups with STAI-X2 outputs substituted by STAI-6 outputs *Positive path coefficients (green arrows) indicate that an increase in the source variable leads to an increase in the dependent variable. Conversely, negative coefficients (red arrows) imply that an increase in the source variable results in a decrease in the dependent variable. For dichotomous variables like toxoplasmosis and borreliosis, positive coefficients indicate that infection is associated with an increase in the dependent variable, while negative coefficients indicate that infection is associated with a decrease in the dependent variable*. *\* indicates p<0.05; ** indicates p<0.01; *** indicates p<0.001*

**Supplementary Table S1.**
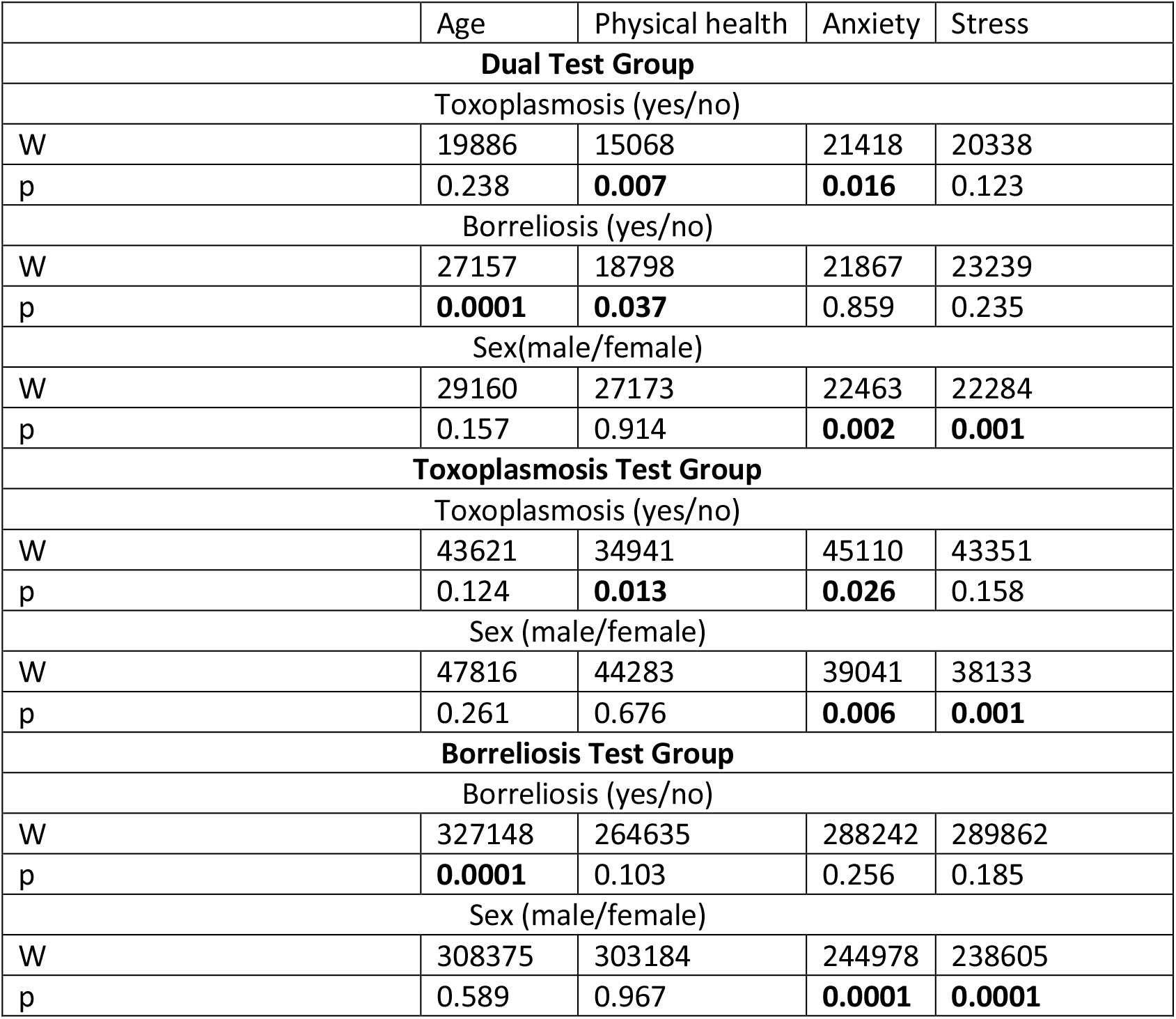
: Post hoc exploration of MANCOVA and ANCOVA results using univariate nonparametric tests *The table presents the Wilcoxon rank-sum test statistics (w) and significance levels (p). Results significant at p < 0.05 are highlighted in bold. For more accurate results, refer to Table 4, which displays the outcomes of multivariate nonparametric tests (partial Kendall correlation test controlled for sex and age, or sex, age, and physical health)*.

